# The evolution of COVID-19 in Italy after the spring of 2020: an unpredicted summer respite followed by a second wave

**DOI:** 10.1101/2020.08.29.20184127

**Authors:** Giuseppe De Natale, Lorenzo De Natale, Claudia Troise, Vito Marchitelli, Antonio Coviello, Karen G. Holmberg, Renato Somma

## Abstract

The coronavirus (COVID-19) pandemic was particularly invasive in Italy during the period between March and late April 2020 then decreased in both in the number of infections and in the seriousness of the illness throughout the summer of 2020. In this discussion, we measure the severity of the disease by the ratio of Intensive Care Units (ICU) spaces occupied by COVID-19 patients and the number of Active Cases (AC) each month from April to October 2020. We also use the ratio of the number of Deaths (D) to the number of Active Cases. What clearly emerges, from rigorous statistical analysis, is a progressive decrease in both ratios until August, indicating progressive mitigation of the disease. This is particularly evident when comparing March-April with July-August; during the summer period the two ratios became roughly 18 times lower. We test such sharp decreases against possible bias in counting active cases and we confirm their statistical significance. We then interpret such evidence in terms of the well-known seasonality of the human immune system and the virus-inactivating effect of stronger UV rays in the summer. Both ratios, however, increased again in October as ICU/AC began to increase in September 2020. These ratios and the exponential growth of infections in October indicate that the virus - if not contained by strict measures - will lead to unsustainable challenges for the Italian health system in the winter of 2020-2021.

## 1. Introduction

COVID-19 had devastating effects in the months of March-May 2020 in Europe. The CFR (Case Fatality Ratio) in European countries (updated on May 26, 2020) reached peaks close to 19% in France, about 16% in Belgium, and around 14% in Italy, UK and Hungary [1, 2]. In this paper, we discuss the reasons for the high CFR in Italy and how these results could potentially be applied to other European countries with very high CFR rates. The main cause, recently confirmed by widespread randomized serological tests in Italy, was a gross underestimation of the true number of infections during the peak of the pandemic. Recent studies indicate that the more accurate number of infected people in Italy was around 1.5 million people, i.e. about six times the tested confirmed cases [3]. With such a correction, the Infection Fatality Rate (IFR), which represents the true lethality of this infection, drops to about 2.3. As occurred in Italy, it is likely that other European countries experienced a higher infection rate than was understood at the time. Another problem, experienced in the most severely impacted Italian region of Lombardy, was the near-collapse of the health infrastructure accompanied by crisis management errors during the infection peak [2]. Lombardy had the highest CFR in Italy, at close to 20%. Other countries with very high CFR likely had similar contexts. It is clearly evidenced that countries with a very strong health system, like Germany, were characterised by much lower CFR [1]. Starting in May 2020, COVID-19 seemed to lose much of its severity in Italy. This was evident to medical staff through direct experience with patients in the main hospitals and prompted active debates in Italy that were reported in Italian media and international press agencies [4]. Such limited, clinical observations raised political-social debates over the necessity of continued, strict containment measures. The evolution of the infection transfers and consequent illnesses during the summer and after the relaxation of the lockdown and other containment measures were far milder than expected by epidemiological forecasts [5,6]. In this paper, we statistically analyse data of ICU occupancy rates and deaths due to COVID-19 as related to the number of active cases from the end of March to October 2020. When rigorously tested, the ratio of ICU occupation to active cases and the ratio of deaths to active cases show significant shifts, thus indicating a change in the evolution of the illness from spring to summer 2020. The likely implications of such changes are then interpreted and discussed, taking into account the possible factors affecting the disease: weakening of the virus, counting bias, and seasonal effects. The results and interpretations are then discussed in light of a possible forecast of what kind of evolution we could expect in the coming months (Autumn-Winter 2020-2021). Finally, we discuss the implications of these observations in the Italian context for the larger global context.

## 2. Data analysis

We study the global, clinical evolution of COVID-19 in Italy using data for ICU occupation numbers, deaths, and active cases in different periods. As [2] point out, ICU numbers and deaths are rather robust data whereas recording of active cases can be strongly biased by heterogeneous testing practices, loss of asymptomatic cases, etc. Recently, the first results of a massive testing campaign to randomly check the percentage of people expressing antibodies to SARS-CoV-2 were released [3]. Such tests confirm, as first hypothesized by [2], that the number of infected people was about six times larger than indicated by official tests, reaching about 1.5 million people (instead of about 250,000 officially tested positive). In the following analyses, we first assume that such high underestimation of active cases has been almost constant during the analysed period; then, we test our results with respect to the maximum bias implied by such an underestimation. Data on active cases, ICU occupation numbers, and deaths in Italy are from the Department of Italian Civil Protection Repository [8]. Here, we report the time evolution of the ratio between the number of people in ICU and the total number of ‘active’ cases (i.e., total less recovered and deaths, at that time), indicated by ICU/AC.

In order to make our estimates more robust, we choose to consider another important and robust indicator: the number of deaths. We therefore also use the ratio between the number of deaths and the number of active cases, indicated as D/AC. In order to get more accurate estimates, we also considered the average time lag from COVID-19 confirmation (the actual data on active infection we have) and the ICU admission as from COVID-19 confirmation to death. According to Wilson et al. (2020)[9], the average time lag between infection confirmation and ICU admission is 6 days whereas the average time lag between infection confirmation and death is 13 days. For this reason, we shift the median day in which we compute the active cases to 6 days before the median day of the ICU number computation; we accordingly shift the median day to compute the number of deaths to 7 days after the ICU median day (so that there are 13 days between the days of active cases computation and the corresponding days of deaths computation). As April 3, 2020 was the day of maximum ICU occupation in Italy for COVID-19 cases we used the third day of each month, from April to October, as the median day to consider ICU occupation numbers. The median days for considering active cases and deaths are chosen accordingly with the described shifts: so that for active cases we take the 27th or 28th of the month before (depending if it has 30 or 31 days), and for deaths we take the 10th of each month considered. In order to obtain more robust estimates of the various data, we choose to average the data during 7 days around each median day (considering also 3 days before and 3 days after the median day). We then computed the two quantities - ICU/AC and D/AC - as the respective averages for each month. The results, for months from April to October, are reported in Table1 and Figure1.

**Table 1.** Ratios ICU/AC and D/AC. Indicated uncertainties are statistical errors computed at 95% probability level (2σ).

|  | **Ratio ICU/AC** | **Ratio D/AC** |
| --- | --- | --- |
| **April** | 0.0593 +/- 0.0068 | 0.0083 +/- 0.0012 |
| **May** | 0.0144 +/- 0.0008 | 0.0019 +/- 0.0008 |
| **June** | 0.0076 +/- 0.0016 | 0.0013 +/- 0.0002 |
| **July** | 0.0046 +/- 0.0004 | 0.0009 +/- 0.0001 |
| **August** | 0.00334 +/- 0.00006 | 0.0005 +/- 0.0002 |
| **September** | 0.0049+/-0.0006 | 0.00042+0.0001 |
| **October** | 0.0061+/-0.0003 | 0.00063+/-0.0001 |

**Fig.1.**
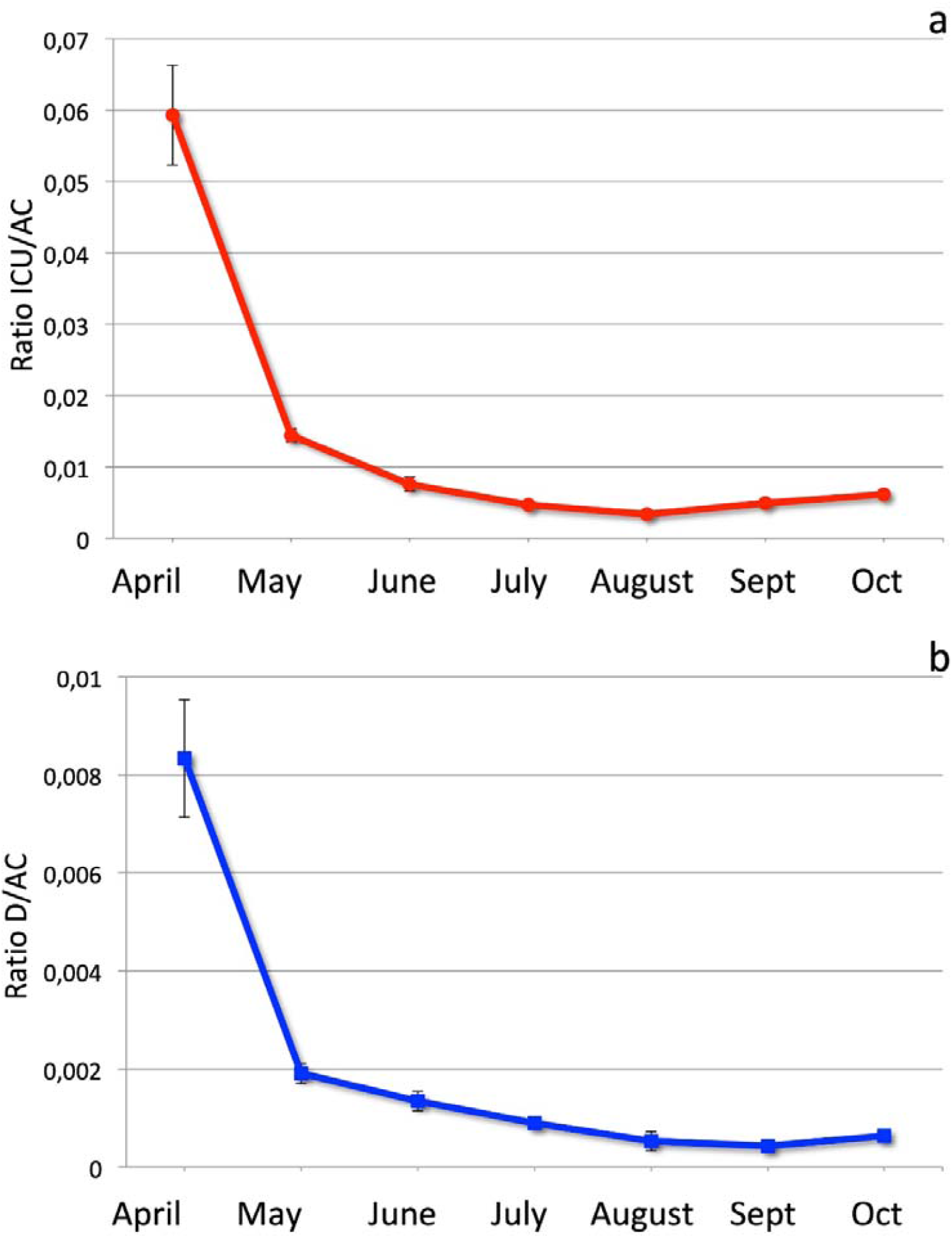
**a)** Ratio between ICU occupation and active cases in each month, from April to October. Error bars are as in table 1 (statistical errors at 95% probability level, i.e. two standard deviations). **b)** Ratio between deaths and active cases in each month, from April to October. Error bars are as in table 1 (statistical errors at 95% probability level, i.e. two standard deviations).

The data in Figure 1 show the statistical uncertainties at 95% (2 standard deviations) and indicate that both the ratios sharply decrease in May, and then progressively continue to decrease in the summer months, with decreasing values clearly separated, well above the statistical uncertainty. Another way to look at such change is to consider the ratios of ICU/AC and D/AC computed for April to each one of the following months.

The computed values of such ratios are reported in Table 2, and shown in Fig.2.

**Table 2.** Relative ratios ICU/AC and D/AC between April and following months. Indicated uncertainties are statistical errors computed at 95% probability level (2σ).

|  | **Relative Ratio ICU/AC** | **Relative Ratio D/AC** |
| --- | --- | --- |
| **April/May** | 4.11 +/- 0.54 | 4.37 +/- 0.84 |
| **April/June** | 7.8 +/- 1.4 | 6.2 +/- 1.3 |
| **April/July** | 12.7 +/- 1.9 | 9.3 +/- 1.8 |
| **April/August** | 17.8 +/- 2.1 | 15.8 +/- 3.5 |
| **April/September** | 12.0+/-2.0 | 19.7+/-4.0 |
| **April/October** | 9.7+/-1.2 | 13.2+/-3.0 |

**Fig.2.**
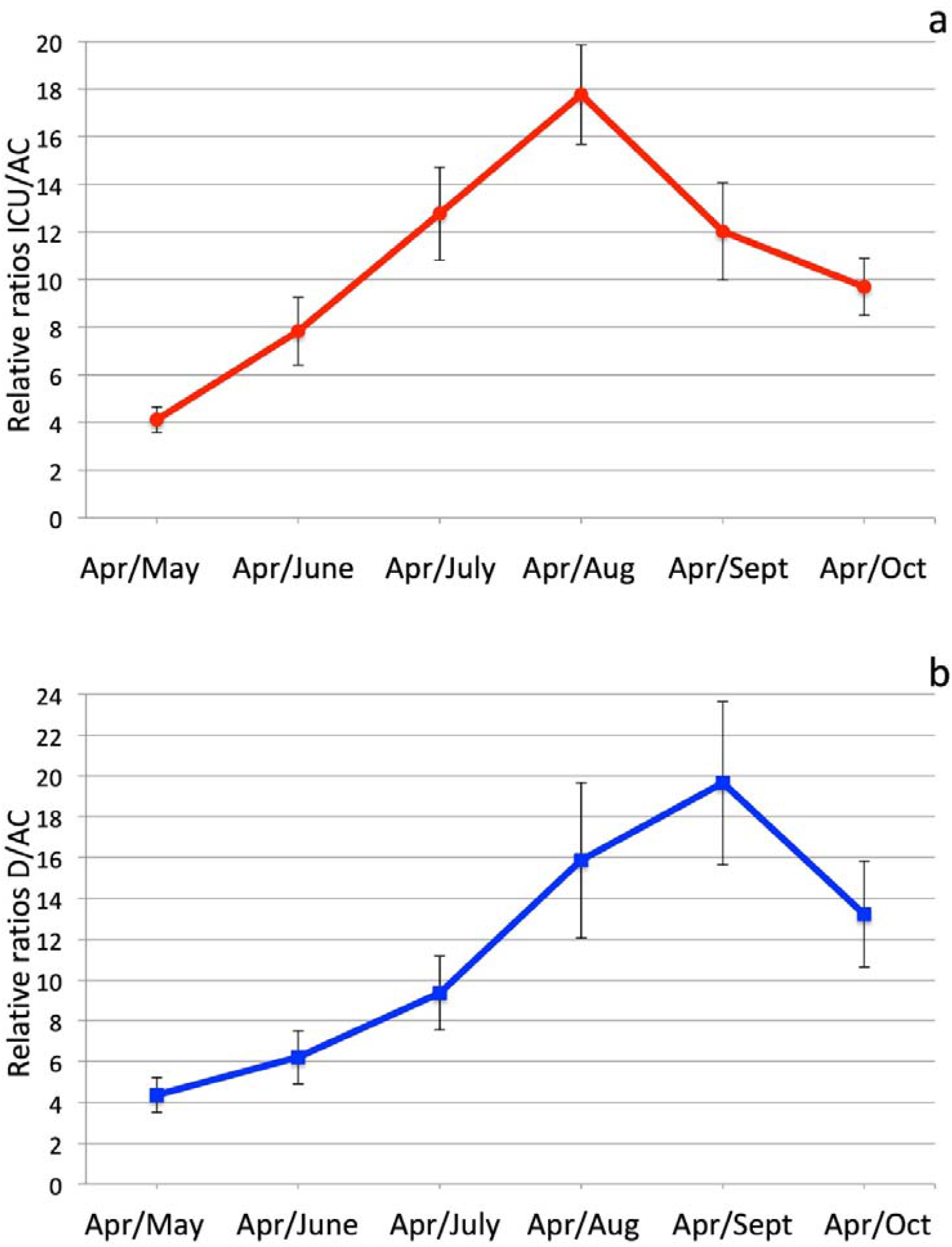
**a)** Relative ratios of ICU/AC in April, with respect to the following months May to August. Error bars are as in table 1 (statistical errors at 95% probability level, i.e. two standard deviations). b) Relative ratios of D/AC in April, with respect to the following months May to August. Error bars are as in table 1 (statistical errors at 95% probability level, i.e. two standard deviations).

**Fig.3.**
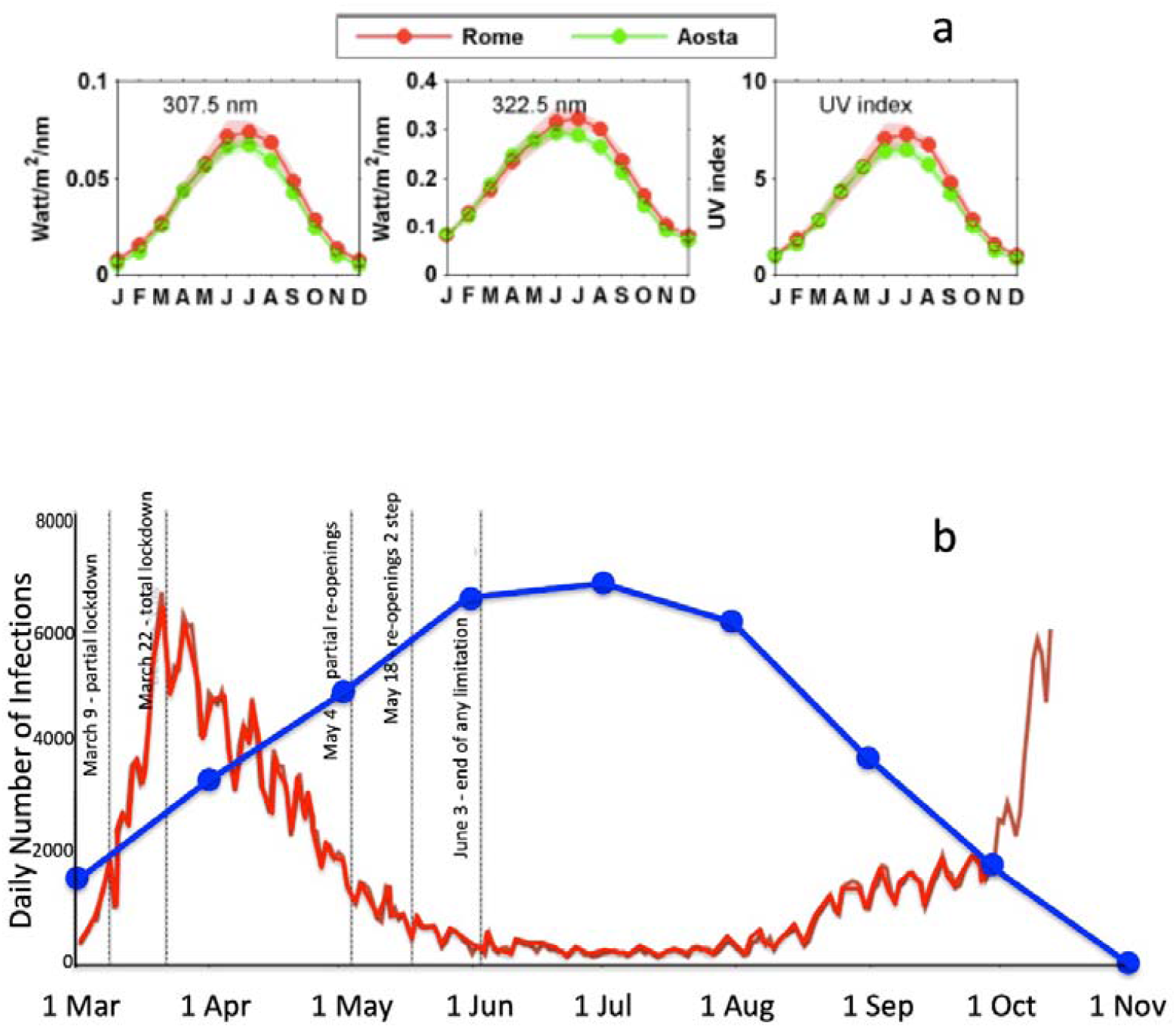
**a)** From left to right: climatologies of the irradiance at 307.5 nm, 322.5 nm, and the UV index at two stations in Italy: Aosta, Northern Italy, Rome, Central Italy, in the period 2006–2015. Shaded envelopes correspond to the standard deviation of the climatological values and the average ratios (redrawn from [21]). **b)** Comparison of the official daily infection curve in Italy from March to mid-October (red line) with the average (2006-2015) UV intensity curve recorded at Rome station (as shown in a), in the period March to November. Note the clear anti-correlation of the two curves, with minimum infection corresponding with maximum UV intensity in the summer months.

**Fig.4.**
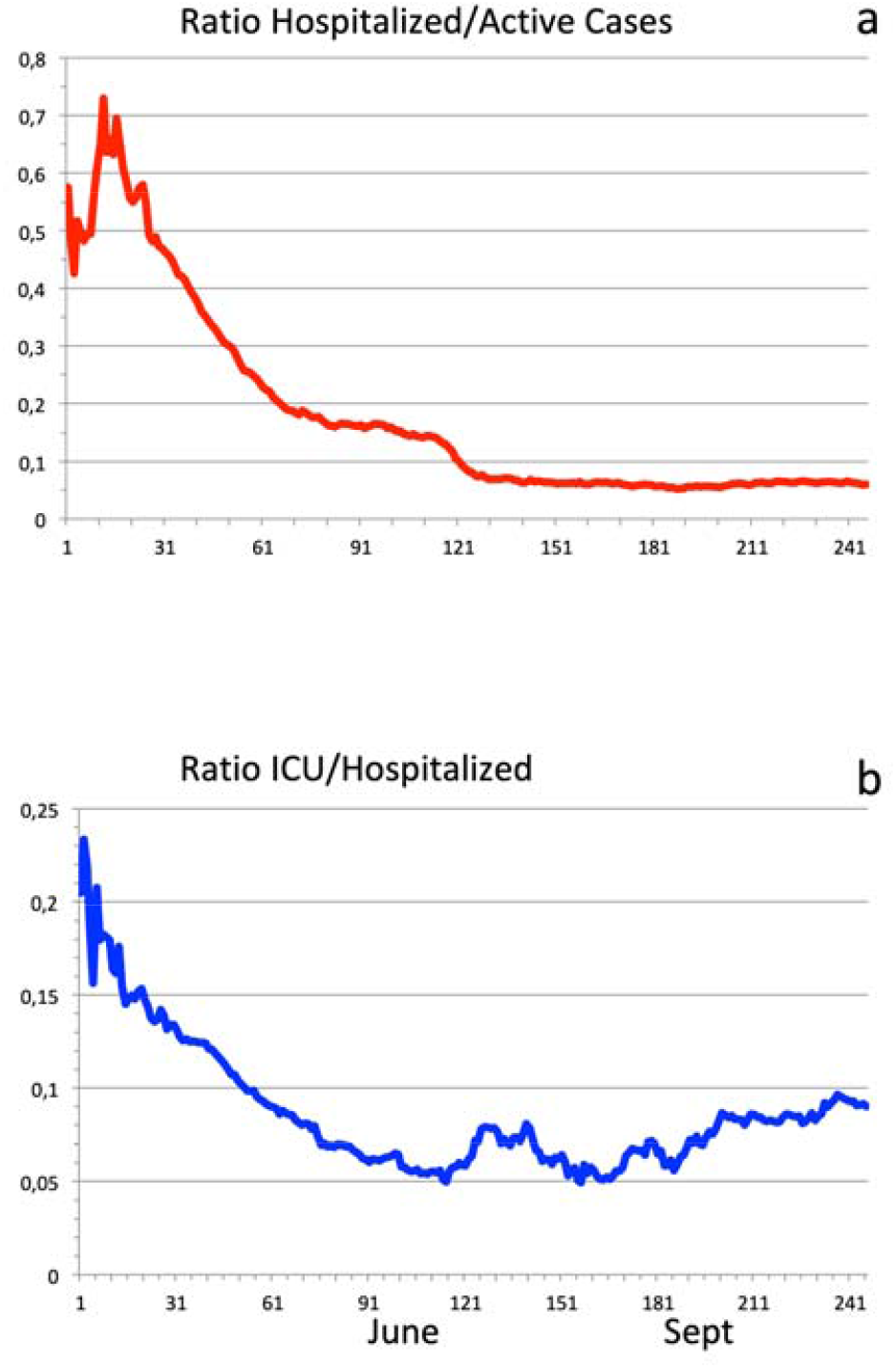
**a)** Ratio between hospitalized Covid-19 patients and total number of active cases, in Italy; b) ratio between ICU occupation and hospitalized patients. The considered period is February 24^th^-October 23^rd^.

**Fig.4.**
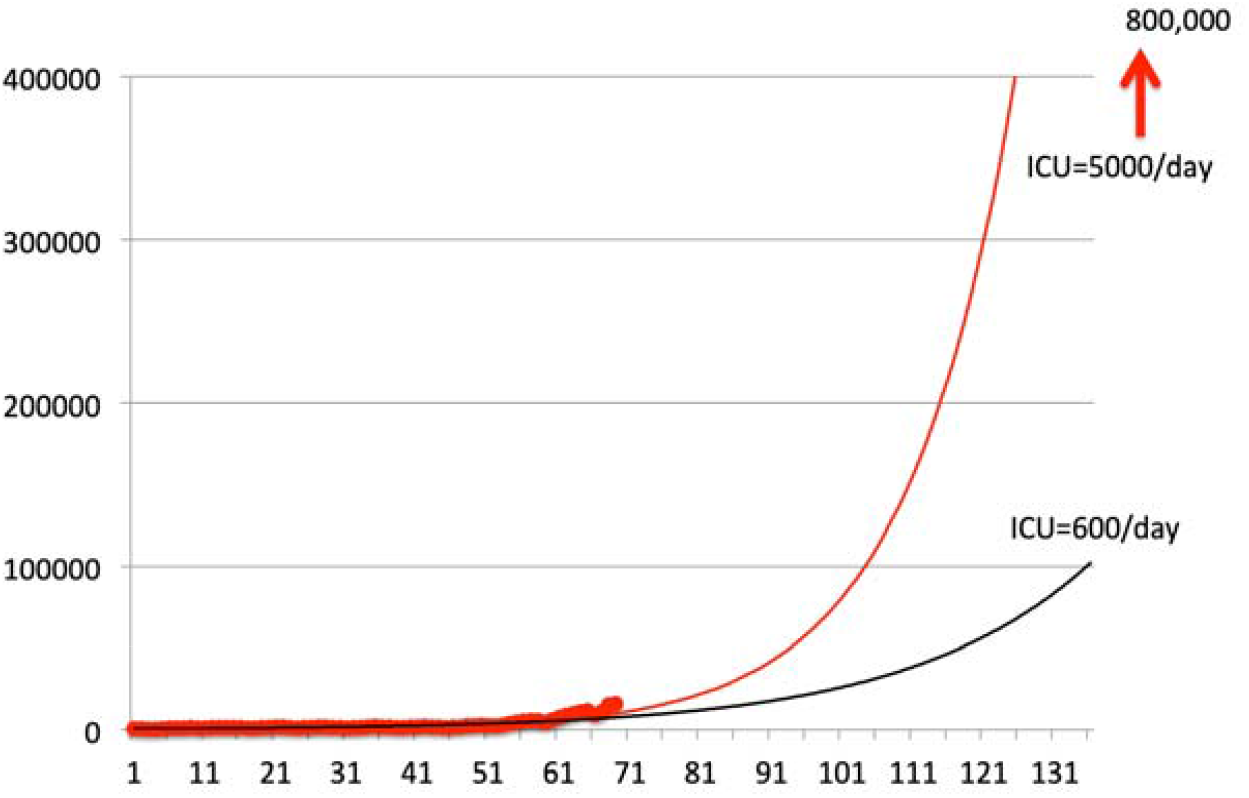
Extrapolated exponential curves for daily infection data since August 15th to October 23^rd^ (red circles). The best fitting exponential curve for all data is shown by the black line; red line shows the best fitting curve using only data since September 15^th^ to October 23^rd^.

As is clear, both ratios progressively and rapidly increase towards and during the summer months: from April to June, the relative ratio (ICU/CA)April/(ICU/CA)June increases of a factor 7.9, and the relative ratios (D/CA)April/(D/CA)June of a factor 6.2; from April to August, the relative ratio (ICU/CA)April/(ICU/CA)August increases of a factor 17.6 and the relative ratio (D/CA)April/(D/CA)August of a factor 16.7.

These increases are exceptionally marked. Assuming the problems of estimating the true number of active cases are constant in time they unequivocally indicate that the illness became progressively milder during the summer months. The next step, however, is to consider the maximum possible bias due to heterogeneity in the counting procedures of infected people. As we said, we are now aware of the massive underestimation of infection rates that occurred in Italy in the first months of the pandemic; serological tests indicated six times more people infected than officially tested positive [3]. Then, in order to compute the maximum bias such a fluctuating underestimation could imply in our computations we assume that all of the underestimation occurred in the months of March-April and that in the last summer months we succeeded in testing all COVID-19 positive cases. Such an assumption divides the March-April ratios (ICU/AC and D/AC) by a factor six to test the obtained values against the values computed for August (with data from July-August).

We then performed a rigorous test of hypothesis to determine if the observed increase of the ratios in August, with respect to the ratios in April though decreased of a factor 6, is significant. We applied the well-known Student’s test [9].

The Student’s test starts by assuming the following formula for the t-variable [9]:

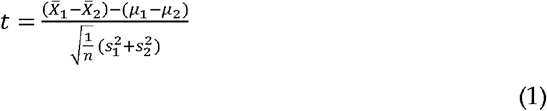

where: 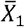 and 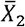 are the average values of ICU/AC (or D/AC) for April and August respectively, and 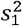 and 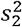 are the respective variances as computed from the samples, n is the number of samples. The number of degrees of freedom is 14-2=12 (14 is the cumulative number of the two samples tested). In the assumption of the null hypothesis, *µ*_1_ and *µ*_2_, the true average values of the two samples are equal, and the value of the t-variable can be tested with respect to the Student’s distribution. For our data, the values result t=9.2 for the ratios ICU/AC, and t=6.3 for the ratios D/AC, which are both largely out of 99.99% probability limits for the Student’s distribution, out of the smallest limits shown in the tables. So, we can very confidently assess that the decrease of the ratios ICU/AC and D/AC, from April to August, is significant for both the variables, even if the maximum bias due to infection counting procedures is assumed, with less than 0.01% probability of being wrong.

After the summer months, however, both the ICU/AC and the D/AC ratios increase again. The minimum value of the first ratio, ICU/AC=0.00334, has been reached in August. In September, the ratio started to increase again, with a value ICU/AC=0.0049; in October, the increase continued, reaching a value ICU/AC=0.0061. A similar increase after the summer is shown by the ratio D/AC; differently from ICU/AC, however, the minimum value D/AC=0.00042 has been reached in September, whereas in October the ratio increased to D/AC=0.00063.

Such an increase of both the indicators in October is well shown by the decrease of the relative ratios April/October; the relative ICU/AC ratio from April to October decreased to 9.7, after in September it had already decreased to 12.0. The relative D/AC ratio from April to October decreased to 13.2, after in September it had reached the maximum value of 19.7.

Following this period, both indicators of the seriousness of the disease markedly decreased starting in April 2020, reaching minimum values at the mid-end Summer (August for ICU/AC, September for D/AC). Both then show increasing values in October 2020, with ICU/AC already increased in September with respect to August. The number of deaths more sharply decreased, however, reaching a minimum value in September, that was roughly 20 times smaller than in April.

## 3. Discussion of results

### 3.1 The summer mitigation

Interpreting the very clear evidence that the disease became progressively less severe, particularly in July-August, is not as simple as it would appear. The simplest explanation could be that the virus itself, which is continuously mutating and adapting to the host [10], lost much of its lethality. With COVID-19, however, we should consider that the severity of the disease is mostly determined by the response of the human immune system [11]. The scientific literature currently provides no clear evidence for the virus becoming substantially mutated and less aggressive. There is evidence for some mutations which actually made the disease less severe [12, 13]; however, there is no indication that such mutations currently have a large diffusion (or have increased compared to past months) in order to significantly mitigate the disease. Hence, the hypothesis that the observed mitigation is due to the large diffusion of a significant virus mutation lacks scientific basis. It is well known that infectious diseases [14] as well as auto-immune diseases are characterized by an evident seasonality [15, 16]. The main reason for this appears to be the seasonal variation of the human immune system response, which has been assessed even in terms of gene expression [17]. In particular, during the summer the immune system response is more effective and less inflammatory. It is proven that during European winters the human immune system has a marked pro-inflammatory character, with increased levels of soluble Il-6 receptor and C reactive protein [17]. An inflammatory response by the immune system, with cytokine storms, has been recognised as the main factor leading to lung and/or other organs failure and death [11].

While there is widespread evidence that almost all flu-like epidemics are strongly dampened during the summer this is even more so in the case of COVID-19: in the acute phase, COVID-19 behaves like an autoimmune syndrome and so is particularly sensitive to the seasonality of the immune response [15, 17]. Other researchers previously noted some seasonal/climate effects [18, 19]. In addition, it has been proven the summer sunlight rapidly inactivates Sars-Cov-2 [20].

The very striking mitigating effect of the summer is easily apparent, although in a partial way considering only the number of infections, by Fig.3, which shows the very clear anti-correlation between the daily number of infection and the intensity of the ultra-violet sunlight (maximum in summer months) from March to mid-October 2020.

The figure shows data on UV radiation intensity, as recorded at two Italian stations: one in northern Italy (Aosta), the other, reported also in Fig.3b in comparison with the daily infection curves, in Rome, central Italy.

It is important to highlight that other explanations, based on the possible bias due to inhomogeneous counting and/or mean age of the infected people in the different periods, as often claimed also on the basis of a relatively younger age inferred for recently recorded infections [22], do not appear to be sufficient to explain the significant decrease of the ICU/AC and D/AC in the summer months. In addition to our demonstration relative to the possible bias of inhomogeneous counting, there is compelling evidence [23] that susceptibility to the virus for individuals older than 20 years is more than twice that of individuals who are younger. Such evidence makes it very unlikely that only very young people in Italy were infected in the summer months and then they did not spread the infection to the older members of the population. The most likely explanation of the inferred lower mean age of infections is that - starting in the summer months - due to the large increase of tests with respect to March-April, mostly asymptomatic cases are recorded [24], whereas in March-April only symptomatic people (comprised mainly of older individuals) were tested. Then, a very strong decrease of the seriousness of the disease in the summer months appears the only realistic explanation of data. The inference that appears is also significant due to the large disparity in the number of tests between the different periods. This is corroborated by the observed increase of the ICU/AC and D/AC after the summer, as will be discussed in the following.

### 3.2 The second infection wave: where are we going?

After the summer months, the two ratios: ICU/AC and D/AC started to increase again. The ICU/AC ratio started to rise again in September and in October reached a value (0.0061) only slightly lower than June (0.0075) but higher than September, August, and July. The D/AC ratio, on the other hands, rose again only in October, reaching a value D/AC=0.00063, higher than September and August, but lower than July.

It is important to stress that this increase of the two ratios after the summer further validates the significance of the summer minima. The number of tests in Italy increased progressively from only a few thousand in March to 160,000-180,000 [8, 25]. It is not possible to imagine a bias, due to the number of tests, capable of explaining the new increase of the two ratios in October or in September and October.

The conclusion that the epidemic was strongly mitigated by the summer appears very robust. What we have described in the Italian case seems to also be able to explain the comparatively low lethality of the virus in recent months observed even in countries experiencing a very large number of infections. Countries like the USA, India, Brazil, and almost all the South American and North African countries, in which epidemics are active, are experiencing much lower CFR (Case Fatality Rates) compared to what European countries experienced in the spring of 2020 [1]. The rates in Israel (CFR=0.8), Arab Emirates (CFR=0.38), or Qatar (CFR=0.17) are extremely low (John Hopkins University, 2020 [26]). Except in a small number of cases, such as the forementioned, it is not probable that non-European countries are recording the real number of infection cases with any higher precision than the European countries. Our conclusions also validate previous observations of COVID-19 worldwide that its evolution was better mitigated in countries characterized by milder climate [18, 19, 26, 27].

The largely mitigated effect of COVID-19 epidemics in Italy during the summer, despite the acute situation in the months of March-April [2], also strongly validates the Italian epidemic management and lockdown strategy. We should note that just after the first reopening - in mid-May 2020 - epidemiological studies predicted a very large increase in the number of infections, severe cases, and deaths [5, 6]. Instead, despite the complete re-opening implemented on June 3, 2020, such dire forecasts were proven inaccurate, confirming the appropriateness of the response and progression from complete lockdown to gradual and then full re-opening.

A further confirmation of the summer mitigation effect comes from the observation of the time behavior of the ICU and Hospitalizations, from late February to late October (Fig.4). In Fig.4a a strong time decrease of the ratio between hospitalized people and active cases is evident, reaching the value, at the end of October, of about 6%. Fig.4b shows the ratio between ICU and hospitalized people, which shows a very clear minimum from the start of June to the start of September; in September and October, this ratio is approaching 10%, from a minimum value of 5%. So, the actual ratio of ICU versus active cases is around 0.6%.

However, the clear evidence for the summer mitigation also rises strong concerns about what we could expect in the winter months. Fig.5 shows the trend of daily tested cases from August 15th to October 23^rd^, 2020. A very sharp increase is evident in the last weeks of that period. The exponential extrapolations, ending at December 31st, were respectively computed as the best fit values for the whole period (black curve) and for the period September 15th to October 23rd (red curve).

Both curves show dramatically high forecasts for the future months. In the best case, the number of infections could rise to up to 100,000 per day; in the worst case, up to 800,000 infections per day could be expected. Such values imply that the corresponding daily death rates could be projected at maximum values of anywhere from 500 to 5000 deaths per day. An estimated 5% of COVID-infected people need hospitalization and about 0.6% require the use of an ICU. This could imply anywhere from 4,000 to 40,000 new hospitalizations per day and 600 to 5000 new ICU occupied per day. The impact of such numbers on the Italian health system is unsustainable.

The actual trend must be decreased as soon as possible to avoid the complete collapse of the hospital system. This requires urgent intensification of public health measures aimed to significantly slow the epidemic.

## 4. Conclusions

We have described how the COVID-19 epidemic in Italy was significantly mitigated during the summer months of 2020. Both the infection rate and the illness criticality appeared suppressed during the summer. This is likely due to the seasonality of immune response, which is more effective and less inflammatory in the summer, combined with the germicidal property of solar UV rays. This effect was frequently misinterpreted by Italian media (and some doctors with direct clinical experience) as a mitigation of the virus and associated illness itself. The subsequent increase in ICU patients and deaths, beginning in September and October, confirm that such a mitigating effect has ended along with the summer. As further confirmation, the daily infection numbers have progressively increased starting from mid-August, with a very sharp acceleration since the end of September. These results potentially explain the very low lethality observed in countries with long lasting summer-like climate but also pose serious concerns for the late Autumn-Winter 2020-2021 in temperate climate zones that experience winters. An exponential progression of the daily infection number, which is indicated by the present data, will lead to unsustainable numbers and the possible collapse of the Italian health system by the end of the winter of 2021. The severity of the COVID crisis is rising in Italy as evidenced by data collected in September-October 2020. Containment of the virus through a curtailing of new infections is vital, pressing, and the only current solution at our disposal until a vaccine and/or monoclonal antibodies are made widely available.

## Data Availability

All data used are in public repository, both Italian and International

